# Gut microbiome and plasma metabolic signatures of one-carbon metabolism differentiate oedematous and non-oedematous severe acute malnutrition

**DOI:** 10.1101/2025.11.26.25341054

**Authors:** Ruairi C. Robertson, Thaddeus J. Edens, Iman Baharmand, Natallia Varankovich, Cherlynn Dumbura, Madhuriben Panchal, Kuda Mutasa, Robert Ntozini, Bernard Chasekwa, Simutanyi Mwakamui, Freddy Francis, Ellen Besa, Thompson Runodamoto, Deophine Ngosa, Florence D. Majo, Kanekwa Zyambo, Claire D. Bourke, Sandra Rukobo, Margaret Govha, Beatrice Amadi, Kusum Nathoo, Mutsa Bwakura-Dangarembizi, Jonathan R. Swann, Paul Kelly, Amee R. Manges, Andrew J. Prendergast, the HOPE-SAM study team

**Affiliations:** Blizard Institute, Queen Mary University of London, London, UK; Zvitambo Institute for Maternal and Child Health Research, Harare, Zimbabwe; School of Population and Public Health, University of British Columbia, Vancouver, Canada; British Columbia Centre for Disease Control, Vancouver, British Columbia, Canada; School of Human Development and Health, Faculty of Medicine, University of Southampton, UK; Tropical Gastroenterology and Nutrition group (TROPGAN), University of Zambia School of Medicine, Lusaka, Zambia; Department of Experimental Medicine, University of British Columbia, Vancouver, BC, Canada; School of Infection and Immunity, University of Glasgow, Glasgow, UK; Department of Paediatrics and Child Health, University of Zimbabwe College of Health Sciences, Harare, Zimbabwe

**Author notes:** **Corresponding author:** Dr. Ruairi Robertson. These authors contributed equally to this work (Amee R. Manges, Andrew J. Prendergast).

## Abstract

Childhood severe acute malnutrition (SAM) occurs as either classical wasting (non-oedematous-SAM; NO-SAM) or with the presence of oedema (oedematous SAM; O-SAM). Both forms of SAM have distinct mortality and nutritional recovery rates, however, the underlying pathophysiology is poorly understood. We collected stool for whole metagenome shotgun sequencing and plasma for metabolic phenotyping from 238 children (79 NO-SAM, 159 O-SAM) hospitalised with complicated SAM and 141 adequately-nourished controls (ANC) from an observational, prospective cohort in Zimbabwe and Zambia. Children with SAM were followed up at 12-, 24- and 48-weeks post-discharge. During hospitalisation, there were significant alterations in taxonomic and functional microbiome diversity between O-SAM and NO-SAM. *Lancefieldella parvula*, a potent producer of hydrogen-sulfide from cysteine, was one of three species significantly elevated in O-SAM versus NO-SAM. Metagenomic analysis showed an overabundance of pathways involved in sulfur amino-acid and one-carbon metabolism in O-SAM compared to NO-SAM. Consistently, the sulfur amino acids cysteine and homocysteine were significantly depleted in the plasma of O-SAM versus NO-SAM, while histidine and various phosphatidylcholines were more abundant. During follow-up, O-SAM and NO-SAM microbiomes remained divergent up to 24 weeks post-discharge whilst alpha diversity recovery was delayed in NO-SAM. Plasma metabolomes also remained significantly different between O-SAM, NO-SAM and ANC throughout 48 weeks of follow-up. These findings demonstrate that distinct microbiome profiles may drive disturbance of systemic one-carbon metabolism in O-SAM and highlight persistent microbiome and metabolic dysfunction during nutritional recovery. This work supports further studies targeting the gut microbiome to correct metabolic disturbances and improve long-term clinical outcomes in complicated SAM.

**One Sentence Summary:** Children hospitalised with oedematous versus non-oedematous severe acute malnutrition have altered gut microbiomes and disrupted one-carbon metabolism, which remain distinct up to 48 weeks post-discharge.

## INTRODUCTION

Severe acute malnutrition (SAM) is the most life-threatening form of undernutrition, affecting at least 12 million children under 5 years worldwide annually (1). Uncomplicated cases of SAM are treated in the community using ready-to-use therapeutic foods. However, SAM often presents with complications including infections, loss of appetite or the presence of nutritional oedema, requiring treatment in hospital. Mortality is as high as 40% in children with complicated SAM during hospitalisation and long-term follow-up (2). SAM is defined as a weight-for-height Z-score (WHZ) ≤ 3, a mid-upper arm circumference <115 mm and/or the presence of bilateral pitting oedema. Children with SAM are therefore classified as either having oedematous-SAM (O-SAM) or non-oedematous SAM (NO-SAM), previously termed kwashiorkor and marasmus, respectively, whilst a small proportion of cases present with both severe wasting and oedema (previously termed marasmic-kwashiorkor). The prevalence of O-SAM versus NO-SAM varies widely across geographical settings (3, 4); mortality rates also differ by region (5). We recently reported on an observational cohort of 745 children with complicated SAM from three hospitals in Zimbabwe and Zambia (Health Outcomes, Pathogenesis and Epidemiology of Severe Acute Malnutrition (HOPE-SAM) study) and found that 64% of children presented with O-SAM, which was associated with a two-fold higher risk of in-patient mortality versus NO-SAM (6). However, post-discharge mortality and hospital readmission were higher, and nutritional recovery was lower, in children with NO-SAM throughout 48 weeks of follow-up (7, 8). Despite these differences in clinical outcomes, World Health Organization protocols for the treatment of SAM do not differentiate between O-SAM and NO-SAM.

The causes of O-SAM have remained enigmatic since the first report of the condition in 1933 (9). Hypotheses of low protein intake, depleted serum albumin and antioxidant status in the development of O-SAM have failed to fully explain its aetiology in both observational studies and randomized trials (10). However, growing evidence supports novel pathways that may differentiate O-SAM from NO-SAM, including differences in lipid and amino acid metabolism (11, 12), reduced expression of heparan sulfate proteoglycan (HSPG) and sulfated glycosaminoglycans (GAG) in the intestine (13). Of recent interest is the association between reduced dietary and circulating sulfur amino acids (methionine, cysteine, homocysteine, and taurine) and O-SAM (14). Many of these sulfur amino acids contribute to one-carbon metabolism, a crucial biochemical pathway involving transfer of methyl groups in DNA methylation and amino acid metabolism. Cysteine is a precursor to the potent antioxidant glutathione which is depleted in O-SAM (15), lending support to the hypothesis that O-SAM is a condition of oxidative stress.

The intestinal tract is a site of complex host-microbiome interactions that may be disturbed in the context of impaired barrier function, enteric infection and inflammation, as commonly observed in SAM (16). Indeed, there is increasing evidence that variation in gut microbial diversity exists between O-SAM and NO-SAM (17, 18). Observational human studies report higher diversity and lower relative abundance of *Prevotellaceae*, *Lachnospiraceae* and *Ruminoccaceae* in the gut microbiome of children hospitalised with O-SAM versus NO-SAM (17, 18). The loss of these oxygen-consuming intestinal anaerobes may contribute to intestinal oxidative stress. Moreover, mechanistic data from animal studies suggest that the gut microbiota of children with SAM may be causally related to wasting as demonstrated through transplantation of stool samples from children with SAM into germ-free mice, resulting in impaired growth and intestinal dysfunction when combined with a deficient diet (19, 20). Furthermore, novel therapeutic diets specifically designed to target the maturation of the early-life gut microbiome can restore growth in mice and in children with acute malnutrition to a greater extent than standard ready-to-use therapeutic foods (RUTF), despite containing lower calorie density than standard RUTF (21, 22). The gut microbiome, therefore, is an essential mediator of nutrient absorption and metabolism in SAM. The mechanisms of this effect, however, are not well defined. Interestingly, data from these experiments show that transfer of SAM microbiomes to germ-free mice results in lower faecal levels of sulfur amino acids methionine and cysteine (20). An undernourished microbiome also contributes to greater susceptibility to intestinal infection, and increased intestinal inflammation and barrier permeability in mice, leading to growth deficits (23, 24).

In this study, we deeply characterized the faecal microbiota and plasma metabolomes of children with complicated O-SAM and NO-SAM during hospitalisation and up to 48 weeks post-discharge, and compared these data to a matched group of adequately-nourished controls, to understand the microbial and metabolic disturbances that differentiate NO-SAM and O-SAM during treatment and convalescence.

## RESULTS

### Baseline characteristics

745 children hospitalised with complicated SAM were recruited to the HOPE-SAM study (25). At enrolment, the median age of children was 17.4 months, 47.7% were female and 21.7% were living with HIV. A subset of these children (n=279) were enrolled in an enteropathy sub-study in which stool, plasma and urine were collected during hospitalisation, at discharge, and at 12-, 24- and 48-weeks post-discharge. For the current analysis, we included 238 children with SAM (159 admitted with O-SAM and 79 admitted with NO-SAM) from the enteropathy sub-study and 141 adequately-nourished controls (ANC) matched for age, sex and HIV status (Fig. S1; see methods for sample selection criteria). ANC were systematically older than SAM children at baseline (29.4 vs 18.1 months respectively; Table 1). The 238 sub-study children with SAM in this analysis had similar baseline characteristics (median age 18.1 months, 48% female, 23% HIV prevalence) to children with SAM who were not included here, many of whom died in hospital (Table S1). In this sub-study, 23 of the 244 children died, with 18 of these deaths occurring in hospital, 12 of which were in children with O-SAM and 6 in children with NO-SAM.

**Table 1.** Baseline characteristics and mortality/readmission outcomes of participants from the HOPE-SAM study included in this analysis.

| Variable | Adequately-nourished children | Children with SAM |  |  |  |
| --- | --- | --- | --- | --- | --- |
|  | Overall | Overall | NO-SAM | O-SAM | *P* value* |
| **Total *n* (%)** | **141** | **238** | **79** | **159** |  |
| **Male, *n* (%)** | 56 (39.7) | 124 (52.1) | 48 (60.8) | 76 (47.8) | 0.081 |
| **Zimbabwe, *n* (%)** | 102 (72.3) | 179 (75.2) | 56 (70.9) | 123 (77.4) | 0.353 |
| **Age, median, months (IQR)** | 29.40 [18.10, 43.80] | 18.10 [13.10, 22.37] | 16.40 [10.95, 21.35] | 18.90 [14.70, 22.50] | **0.011** |
| **WHZ (median, IQR)** | 0.17 [-0.41, 0.82] | -2.82 [-3.92, -1.52] | -3.76 [-4.44, -3.12] | -2.04 [-3.40, -0.96] | **<0.001** |
| **MUAC [mm, median (IQR)]** | 152 [144, 161] | 119 [110, 130] | 113 [105, 121] | 122 [113, 136] | **<0.001** |
| **HAZ (median, IQR)** | -1.71 [-2.74, -0.89] | -2.98 [-3.81, -2.02] | -3.14 [-4.01, -1.98] | -2.86 [-3.73, -2.06] | 0.253 |
| **Birth weight, kg, median (IQR)** | 3.00 [2.70, 3.40] | 2.95 [2.50, 3.30] | 2.60 [2.20, 3.03] | 3.10 [2.70, 3.36] | **<0.001** |
| **Living with HIV, *n* (%)** | 41 (29.1) | 54 (22.7) | 24 (30.4) | 30 (18.9) | 0.067 |
| **Currently breastfed** | 32 (22.7) | 43 (18.1) | 28 (35.4) | 15 (9.4) | **<0.001** |
| **Died** | NA | 23 (9.7) | 9 (11.4) | 14 (8.8) | 0.687 |
| **Readmitted to hospital** | NA | 29 (13.1) | 9 (12.3) | 20 (13.4) | 0.988 |
| **Diarrhea (last 24h)** | NA | 105 (44.1) | 38 (48.1) | 67 (42.1) | 0.463 |

\*NO-SAM versus O-SAM

### Oedema status is associated with significant variation in gut microbiome composition and function during hospitalisation for SAM

Whole metagenome shotgun sequencing was performed on all available stool samples collected from participants (baseline stool samples: 119 admitted with O-SAM, 54 admitted with NO-SAM, 136 ANC). First, we explored factors that contribute to the variation in gut microbiome composition during hospitalisation for complicated SAM, modelling 17 biological and environmental covariates that could plausibly drive microbiome composition (EnvFit analysis - Bray–Curtis dissimilarity). As expected, age explained the greatest amount of variation in gut microbiome composition (R^2^ = 0.136; P < 0.001; Fig. 1a). Only three other covariates, WHZ (R^2^ = 0.054; P = 0.007), oedema status (R^2^ = 0.041; P = 0.007) and current breastfeeding status (R^2^ = 0.029; P = 0.014), were significant correlates of variation in microbiome composition. Number of days of hospitalisation before enrolment, baseline length-for-age Z-score (LAZ), study site and HIV status were the next strongest contributors to microbiome variation but did not meet the significance threshold (p<0.05).

**Figure 1.**
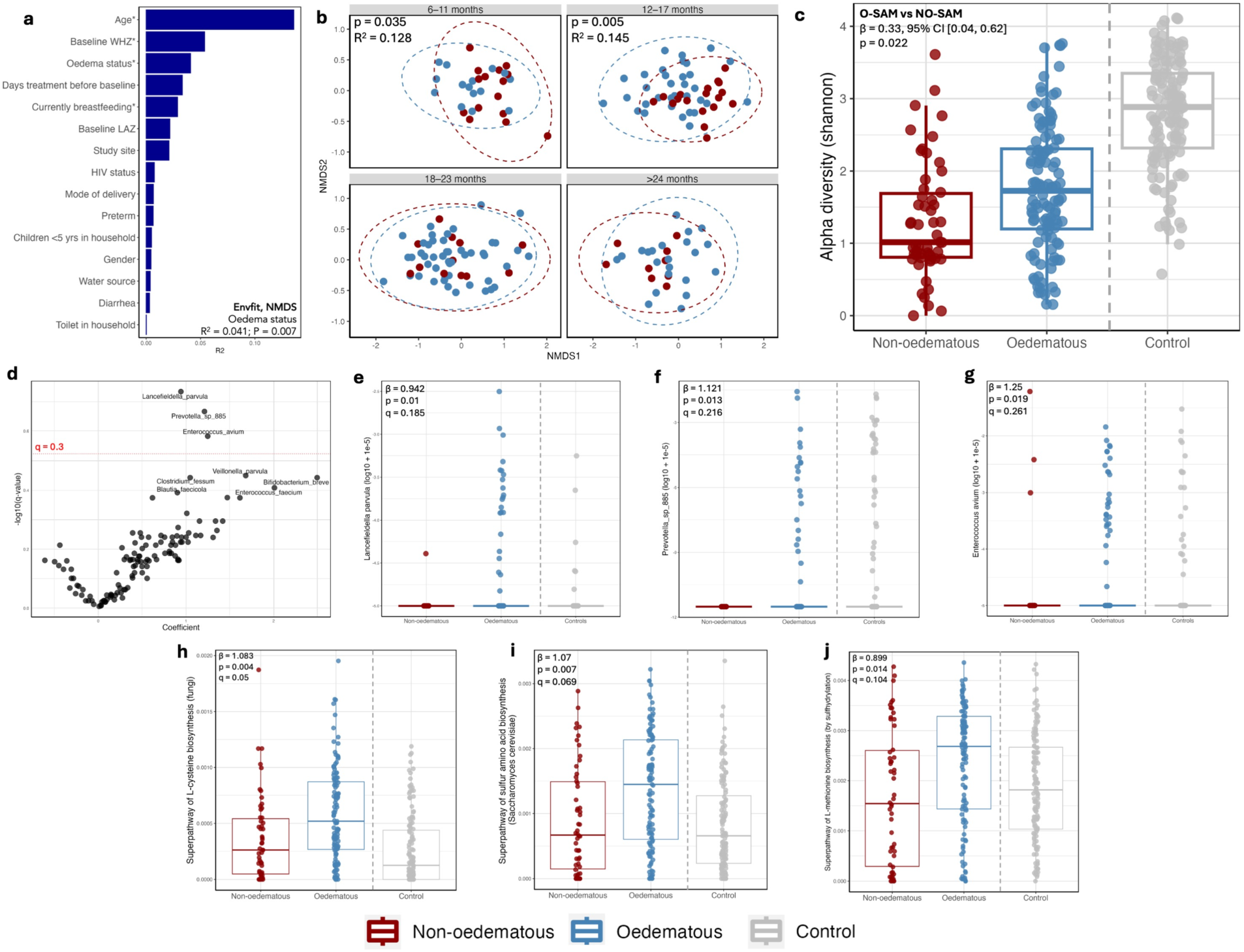
The taxonomic composition of the gut microbiome of children hospitalised for severe acute malnutrition differs by oedema status. (a) Results from EnvFit model (Bray–Curtis dissimilarity) examining baseline characteristics driving variation in species composition of the gut microbiome (*p<0.05). (b) NMDS ordination plots coloured by oedema status with P values and R^2^ values from EnvFit models separated by age brackets. (c) Alpha diversity (Shannon index) with results of multivariable regression model comparing O-SAM versus NO-SAM. ANC included as reference but not included in statistical model. (d) Volcano plot of results from MaAsLin2 regression testing of differential abundance between O-SAM and NO-SAM (positive coefficients indicate higher abundance in O-SAM). Red dotted line indicates false discovery rate threshold for q values indicating significant taxa (e-g) Relative abundances (log10 + 1e-5) and results from MaAsLin2 regression for selected species and (h-j) selected gene pathways (relative abundances).

Due to the strong effect of age on microbiome composition and the wide range of ages in this sample subset of hospitalised children (6 months – 5 years), we also applied EnvFit analysis to sub-categories of ages (6-11 months, 12-17 months, 18-23 months, >24 months). This identified a significant association between oedema status and microbiome composition at age 6-11 months (R^2^ = 0.128, P = 0.035) and 12-17 months (R^2^ = 0.145, P = 0.005) but not in older age groups, suggesting that the association between gut microbiome composition and oedema status was more prominent in early childhood and diminished with age (Fig. 1b). In the analyses of samples from ANC alone, age (R^2^ = 0.387), current breastfeeding status (R^2^ = 0.236), HIV status (R^2^ = 0.063) and household water source (R^2^ = 0.045) explained significant variation in gut microbiome composition.

We replicated these analyses using functional metagenomic pathways to assess the effect of the same biological and environmental variables on gut microbiome functions. Consistently, age explained the biggest variation in microbiome function (R^2^ = 0.08, P = 0.002) but was the only significant feature in the model. However, similar to the microbiome composition results, there was a significant association between oedema status and microbiome function in the 6-11 month age category (R^2^ = 0.192, P = 0.003; Fig. S2a). Oedema also explained significant variation in gut microbiome in the >24 month age category (R^2^ = 0.39, P < 0.001; Fig. S2a). In the analyses of samples from ANC alone, age (R^2^ = 0.278), current breastfeeding status (R^2^ = 0.179), HIV status (R^2^ = 0.131) and study site (R^2^ = 0.069) explained significant variation in gut microbiome metagenomic pathways.

We next compared microbiome alpha diversity between NO-SAM, O-SAM, and ANC (median Shannon diversity: 1.01, 1.73 and 2.89 respectively; Fig. 1c). Using multivariate linear regression, we found a significant association between oedema status and Shannon diversity (β = 0.33, 95% CI [0.04, 0.62], p = 0.022) whereby children with O-SAM displayed significantly higher diversity than children with NO-SAM (Fig. 1c). When including an interaction term in the model, we observed that the association between oedema and Shannon diversity depended on age. The diversity difference was strongest at younger ages, with O-SAM children showing higher diversity, but this difference diminished as age increased (interaction β = –0.036, p = 0.013; Fig. S2b). We also examined gene richness, an indicator of metagenomic diversity, and found similar results; children with O-SAM versus NO-SAM displayed significantly greater metagenomic gene richness (β = 22290, 95% CI [4841, 39740], p = 0.013; Fig. S2c). All children with SAM were on prophylactic antibiotic therapy during hospitalization as part of WHO protocols which limited our ability to compare microbiomes between SAM and ANC as an effect of nutritional status. Both O-SAM and NO-SAM groups had significantly lower Shannon diversity richness than ANC (p <0.001) whereas metagenomic richness was only depleted in NO-SAM, but not O-SAM, versus ANC.

Collectively, these results demonstrate that, during hospitalisation for SAM, children with O-SAM and NO-SAM have distinct microbiome composition and function, particularly at younger ages.

### The gut microbiome of oedematous SAM is enriched in H_2_S-producing species and one-carbon metabolism pathways

We next explored specific taxonomic and functional differences in the gut metagenome between O-SAM and NO-SAM. Using multivariable general linear models adjusted for age, HIV and current breastfeeding status, three species were found to have significantly larger relative abundance in O-SAM versus NO-SAM (Fig. 1d-g; Table S2): *Lancefieldella parvula* (previously classified as *Streptococcus parvulus* and subsequently *Atopobium parvulum;* β = 0.942, p = 0.010, q = 0.185)*, Enterococcus avium* (β = 1.25, p = 0.019, q = 0.261) and *Prevotella sp. 885* (β = 1.121, p = 0.013, q = 0.216). *Veillonella parvula, Bifidobacterium breve* and *Enterococcus faecium* were also elevated in O-SAM versus NO-SAM but did not meet false-discovery rate significance thresholds. *L. parvula* and *V. parvula* are both potent producers of hydrogen sulfide (H_2_S) via the catabolism of cysteine and were also both significantly elevated in O-SAM versus ANC. Indeed, *L. parvula* colonization was rare in ANC children (6/136; 4%) and in children with NO-SAM (1/54; 2%) but colonized 18% (22/119) of children with O-SAM.

We subsequently performed the same analyses with metagenomic pathways and identified 41 significantly differentially abundant pathways between O-SAM and NO-SAM (Table S3), 38 of which were elevated in O-SAM, the most significant of which were those involved in nicotinamide adenine dinucleotide (NAD) salvage (*PYRIDNUCSAL PWY – NAD salvage pathway I PNC VI cycle*). In addition, there were several pathways involved in cysteine, one-carbon and sulfur-amino acid metabolism (*PWY 6293 - superpathway of L-cysteine biosynthesis (fungi); PWY-821 - superpathway of sulfur amino acid biosynthesis (Saccharomyces cerevisiae); PWY 5030 - L histidine degradation III; PWY 5345 - superpathway of L methionine biosynthesis by sulfhydrylation; HSERMETANA PWY - L methionine biosynthesis III; PWY 5028 - L histidine degradation II*; *PWY.6549 – L-glutamine biosynthesis III)* all of which were elevated in O-SAM stool metagenomes (Fig. 1h-j; Table S3). We further examined enzymes involved in cysteine and homocysteine metabolism and found that *S*-adenosylhomocysteine deaminase (EC 3.5.4.28), adenosylhomocysteinase (EC 3.3.1.1) and cystathionine β-lysase (EC 4.4.1.8) were all elevated in O-SAM versus NO-SAM amongst other enzymes (Table S4).

In a subset of children (n=14 NO-SAM, n=31 O-SAM), we conducted metagenomic sequencing on gastric fluid collected from children who had nasogastric tubes in place for in-patient feeding. Gastric pH was very high among children with SAM (38/45 with pH>4 i.e. achlorydia) but did not differ significantly between O-SAM and NO-SAM (Fig. S3a). There was no difference in gastric microbiome alpha diversity between O-SAM and NO-SAM (p=0.38; Fig. S3b), however the relative abundance of *Klebsiella pneumoniae* (β = -2.17, p = 0.014, q = 0.262) and Enterobacter spp. (β = -1.9, p = 0.002, q = 0.142) was significantly lower in O-SAM versus NO-SAM whilst an unclassified species from the *Porphyromonadaceae* family (GGB1088; β = 1.9, p = 0.023, q = 0.29) was significantly elevated in O-SAM (Fig. S3c-e).

Taken together, taxonomic and metagenomic differential abundance data suggested that the gut microbiome of children with O-SAM was overrepresented by a small number of species, including those involved in production of H_2_S from cysteine and genes involved in sulfur amino acid metabolism.

### Cysteine and homocysteine are depleted in the plasma metabolomes of children with O-SAM

We next measured the metabolic phenotypes in a subgroup of 177 children with available plasma samples from enrolment (78 O-SAM, 30 NO-SAM and 69 ANC) using liquid chromatography-mass spectrometry. Plasma metabolic phenotyping was conducted on HIV-negative children enrolled in the two Zimbabwe sites (see methods for sample selection criteria). A significant partial least squares-discriminant analysis (PLS-DA) model was obtained by comparing the O-SAM versus NO-SAM plasma metabolomes (permutation test: Classification error rate (CER) = 0.199, p = 0.001; Fig. 2a). From this model, five phosphatidylcholine metabolites and a ceramide were identified to be greater in the circulation of children with O-SAM versus NO-SAM (Fig. S4a-f), while cysteine was present in reduced amounts (Fig. 2b). Homocysteine, tryptophan and kynurenine were also among the top 50 metabolites contributing to variation in the model and all were lower in O-SAM compared to NO-SAM. Sparse PLS-DA (sPLS-DA) using lasso penalisation confirmed these findings (Table S5). Using linear regression assessing all 294 metabolites individually against oedema status, these 4 metabolites were all significantly depleted in O-SAM compared with NO-SAM (Fig. 2c-I; Table S8): cysteine (log2 fold change: -0.54 [95% CI: -0.79, -0.29]; adjusted p = 0.007), homocysteine (-0.66 [-1.12, -0.2]; adjusted p = 0.02), tryptophan (-0.70 [-1.15, -0.25]; adjusted p = 0.036) and kynurenine (-0.5 [-0.87, -0.14]; adjusted p = 0.058). Cysteine (p = 0.007) and homocysteine (p = 0.031) remained significantly reduced in O-SAM vs NO-SAM following multivariable regression, adjusting for sex, age and current breastfeeding status. Cysteine (p=0.0003) and kynurenine (p=0.007) were also both significantly depleted in O-SAM compared with ANC, whereas tryptophan was significantly depleted in both O-SAM and NO-SAM versus ANC. We also compared these metabolites across grades of oedema severity and found that the greatest depletion occurred in those with the highest degree of oedema (Fig. S5a-d). In contrast, 42 metabolites were significantly elevated in O-SAM versus NO-SAM. These included histidine, trimethylamine-*N*-oxide (TMAO) and glycine, in addition to multiple ceramides and phosphatidylcholine species, all of which remained significant following adjustment for confounders.

**Figure 2.**
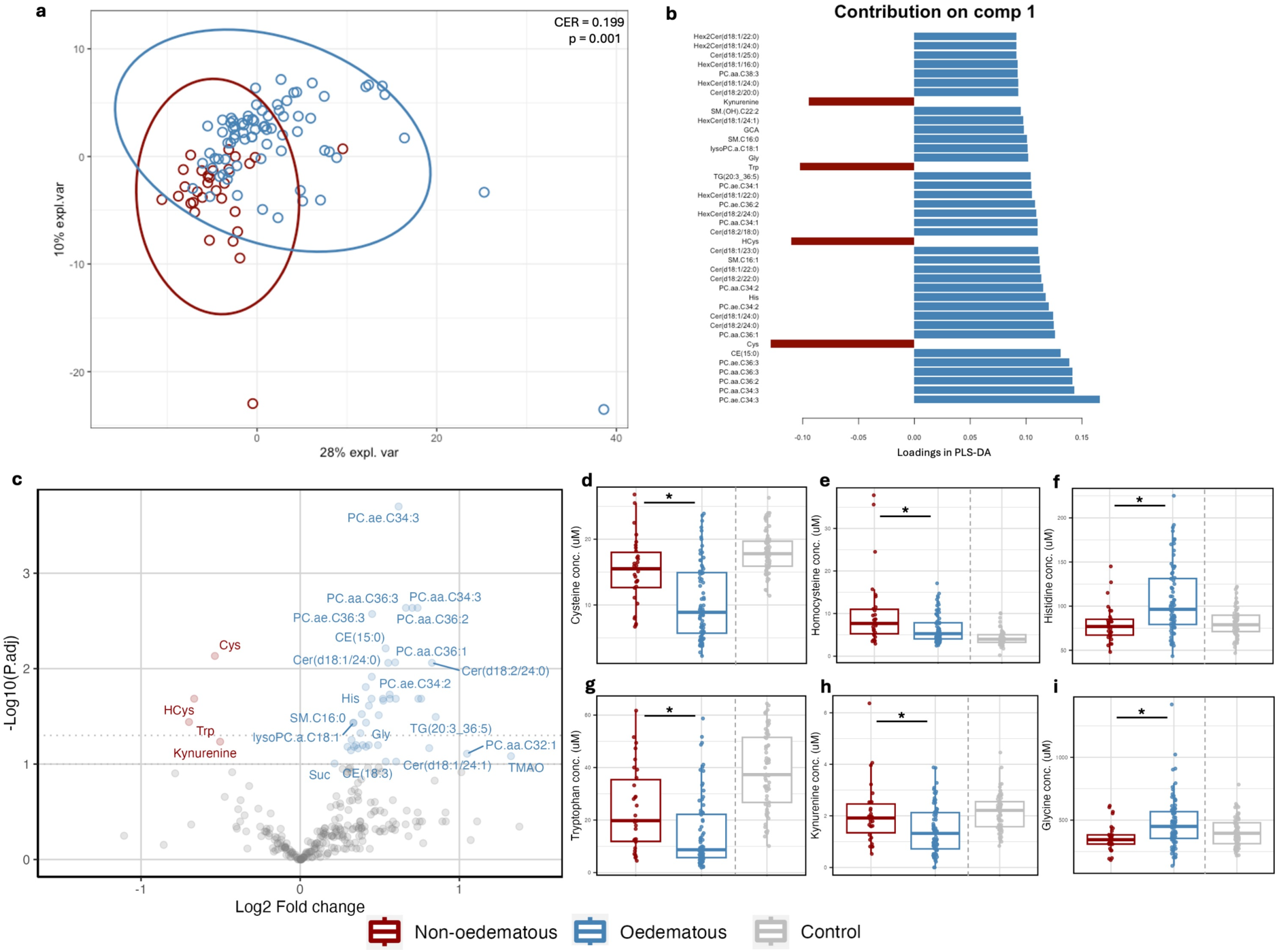
Plasma metabolic phenotypes of children with O-SAM vs NO-SAM during hospitalisation. (a) PLS-DA model of O-SAM and NO-SAM during hospitalisation including model results from permutation test (p value and classification error rate (CER)). (b) Top metabolites contributing to model separation based on model loadings scores. (c) Volcano plot of metabolites significant in univariate linear regression models in O-SAM versus NO-SAM. (d-i) Concentrations of selected metabolites significantly different between O-SAM and NO-SAM (*adjusted p value <0.05 in univariate linear regression). Concentrations of metabolites from ANC are plotted for reference but not included in statistical models.

We explored associations between the metagenomic and metabolomic datasets to identify multi-omic relationships that distinguish O-SAM and NO-SAM by integrating microbiome species, gene pathways and plasma metabolite datasets from 86 participants with paired stool microbiome and plasma metabolome data at baseline (22 SAM, 64 O-SAM). The multi-omic block sPLS-DA model had a lower balanced error rate (BER; 0.107) when classifying O-SAM and NO-SAM compared with sPLS-DA predictions using individual omics datatsets (microbiome species BER – 0.53; microbiome pathways BER – 0.444; plasma metabolites BER – 0.223; Fig. S6a-b). The integrated model identified correlations between several microbiome and metabolomic features (Fig. S6c) and network modelling identified the metagenomic pathways *PWY 5345 - superpathway of L methionine biosynthesis by sulfhydrylation* and *PWY 821 - superpathway of sulfur amino acid biosynthesis (Saccharomyces cerevisiae)* as central nodes in the network (Fig. S6d) suggesting that they are highly connected across omic layers. These two pathways were highly correlated with some phosphatidylcholine metabolites (r >0.8; Fig. S6d) but also exhibited modest correlations with plasma cysteine (r = -0.38 and r = -0.34 respectively) and other metabolites (Table S6a-c).

Collectively, these data highlight distinct metabolic signatures between children with O-SAM and NO-SAM, specifically reduced cysteine and homocysteine and elevated histidine in O-SAM, among other metabolites, which are correlated with changes in gut microbiome composition and function.

### Microbiome and metabolic disturbances persist during convalescence

We subsequently tested whether differences in gut microbiome and plasma metabolomes between O-SAM, NO-SAM and ANC children persisted during one year of follow-up (at 12 weeks, 24 weeks and 48 weeks post-discharge). Beta diversity (Bray–Curtis dissimilarity assessed using permutational analysis of variance) analyses highlighted ongoing significant differences in gut microbiome taxonomic profiles in NO-SAM at 12-weeks post-discharge compared with O-SAM (adjusted p = 0.032, R^2^ = 0.014; Fig. 3a) and compared with ANC (adjusted p = 0.031, R^2^ = 0.009; Fig. S7a) suggesting delayed recovery of taxonomic composition in NO-SAM but not in O-SAM. No differences were apparent between groups at 24- and 48-weeks follow-up (Fig. 3a and Fig. S7b-c).

**Figure 3.**
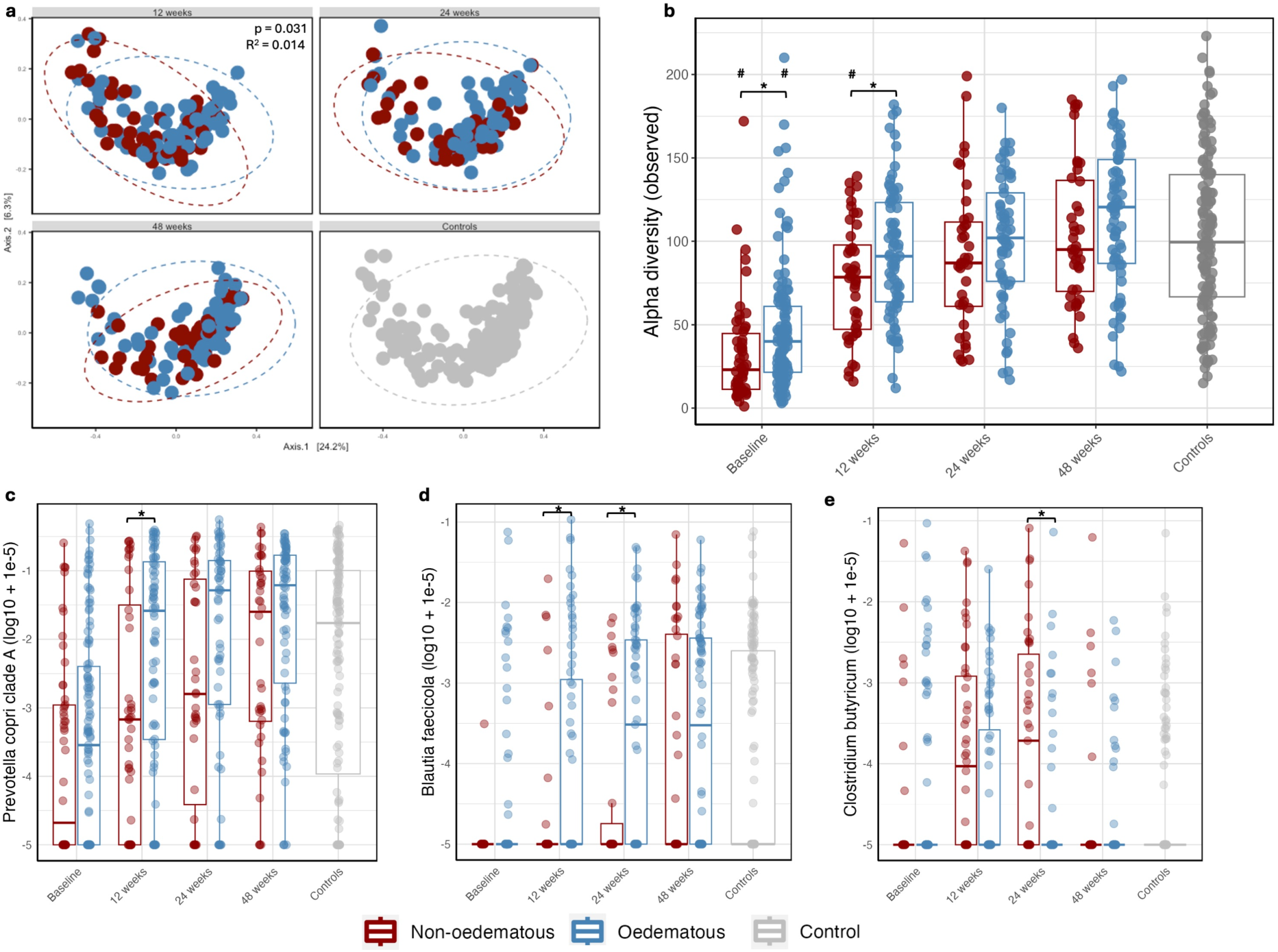
Gut microbiome recovery in children with O-SAM and NO-SAM during 48 weeks of follow-up post-discharge. (a) NMDS ordination and permutational analysis of variance (PERMANOVA) results comparing O-SAM versus NO-SAM species composition with P values and R^2^ values from EnvFit models for each follow-up visit. (b) Shannon diversity in O-SAM and NO-SAM at follow-up visits and ANC (*indicates significant model comparing O-SAM versus NO-SAM within a follow-up visit; ^#^indicates significant result for a model comparing O-SAM or NO-SAM with ANC). (c-e) Relative abundances (log10 + 1e-5) of selected species during follow-up post-discharge in O-SAM versus NO-SAM (*indicates significant model comparing O-SAM versus NO-SAM within a follow-up visit).

Alpha-diversity (taxonomic richness) remained significantly higher in O-SAM vs NO-SAM at 12-weeks post-discharge in univariate analysis (β = 16.315, 95% CI [2.38, 30.25], p = 0.022) but not following adjustment for covariates (Fig. 3b). Taxonomic richness in NO-SAM, but not O-SAM, remained significantly lower versus ANC at 12-weeks post-discharge in multivariable analysis (β = -24.998, 95% CI [-49.62, -0.37], p = 0.047) but not at later visits, suggesting a delayed recovery of microbiome diversity in NO-SAM in the immediate period following hospital discharge. No significant differences were observed in beta-diversity of microbiome gene pathways or gene richness at any follow-up visits.

We further examined species relative abundances at the same follow-up time points. Species that were differentially abundant at baseline between O-SAM and NO-SAM were no longer differentially abundant at follow-up time points except for *Prevotella sp. 885* which was significantly elevated in O-SAM at 24 weeks post-discharge (β = 1.77, p = 0.007, q = 0.199). Children with O-SAM exhibited significantly elevated abundances of various *Blautia* (*B. massiliensis, B. faecicola* at 12 weeks; *B. obeum* and *B. faecicola* at 24 weeks) and *Prevotella* species (*P. copri* clades A, C and G at 12 weeks; *P. copri* clade G, *P. stercorea* at 24 weeks) in addition to significantly elevated abundances of *Ruminococcus torques* at 12 weeks relative to children with NO-SAM (Fig. 3c-e). Conversely, children with O-SAM had significantly reduced relative abundances of *Enterocloster aldenensis* and *Erysipelatoclostridium ramosum* at 12 weeks, *Clostridium butyricum, Akkermansia muciniphila* and *Eggerthella lenta* at 24 weeks and *Clostridium SGB4750* at 48 weeks compared with NO-SAM (Tables S8-10). Most metagenome pathways that differed significantly between O-SAM and NO-SAM at baseline were no longer different at follow-up visits, however *HSERMETANA PWY - L methionine biosynthesis III* continued to show significant differences between groups at weeks 12 and 24 although with varying direction of effect. Few pathways remained different between groups by week 48 (Tables S11-13).

Plasma metabolic phenotypes continued to differ significantly between O-SAM and NO-SAM at the 12 week (permutation test: CER = 0.328, p = 0.038) and 48-week visit (permutation test: CER = 0.311, p = 0.044) in PLS-DA analysis indicating persistent metabolic differences between children with O-SAM and NO-SAM throughout follow-up (Fig. 4a-c). Metabolites driving this separation at follow-up visits included dehydroepiandrosterone sulfate (DHEAS) and tryptophan, which were reduced in O-SAM compared to NO-SAM participants, and choline and betaine which were increased in O-SAM (Fig. 4d-f; Tables S14-16). We also compared the metabolic phenotypes of O-SAM and NO-SAM to the ANC children to assess metabolic “recovery” and observed significantly different metabolic phenotypes in PLS-DA analyses relative to controls in both groups at all follow-up visits (all p <0.005; Fig. S7d-f), suggesting that metabolic dysfunction continued up to a year after discharge from hospital. We applied multivariate regression analysis on one-carbon metabolites that were differentially abundant between O-SAM and NO-SAM at baseline (Fig. 4g-j). Cysteine was no longer significantly different between O-SAM and NO-SAM at any of the follow-up visits. Conversely to baseline, homocysteine was elevated in O-SAM versus NO-SAM at 12 weeks follow up (0.31 [0.07, 0.54]; p = 0.008) but did not significantly differ at subsequent follow-up visits. It was also significantly higher relative to ANC at 48 weeks (p = 0.007). Histidine concentrations no longer differed significantly between O-SAM and NO-SAM at any follow-up visits, although histidine was significantly higher in O-SAM versus ANC at 48 weeks (p = 0.043). Tryptophan remained significantly depleted compared with ANC at 12 weeks and to both NO-SAM and ANC at 24- and 48-weeks. These data suggest that the deficiencies in plasma cysteine observed in O-SAM are short-lived and tend to recover during follow-up whereas disruption to tryptophan and other metabolites persists after hospitalisation for SAM.

**Figure 4.**
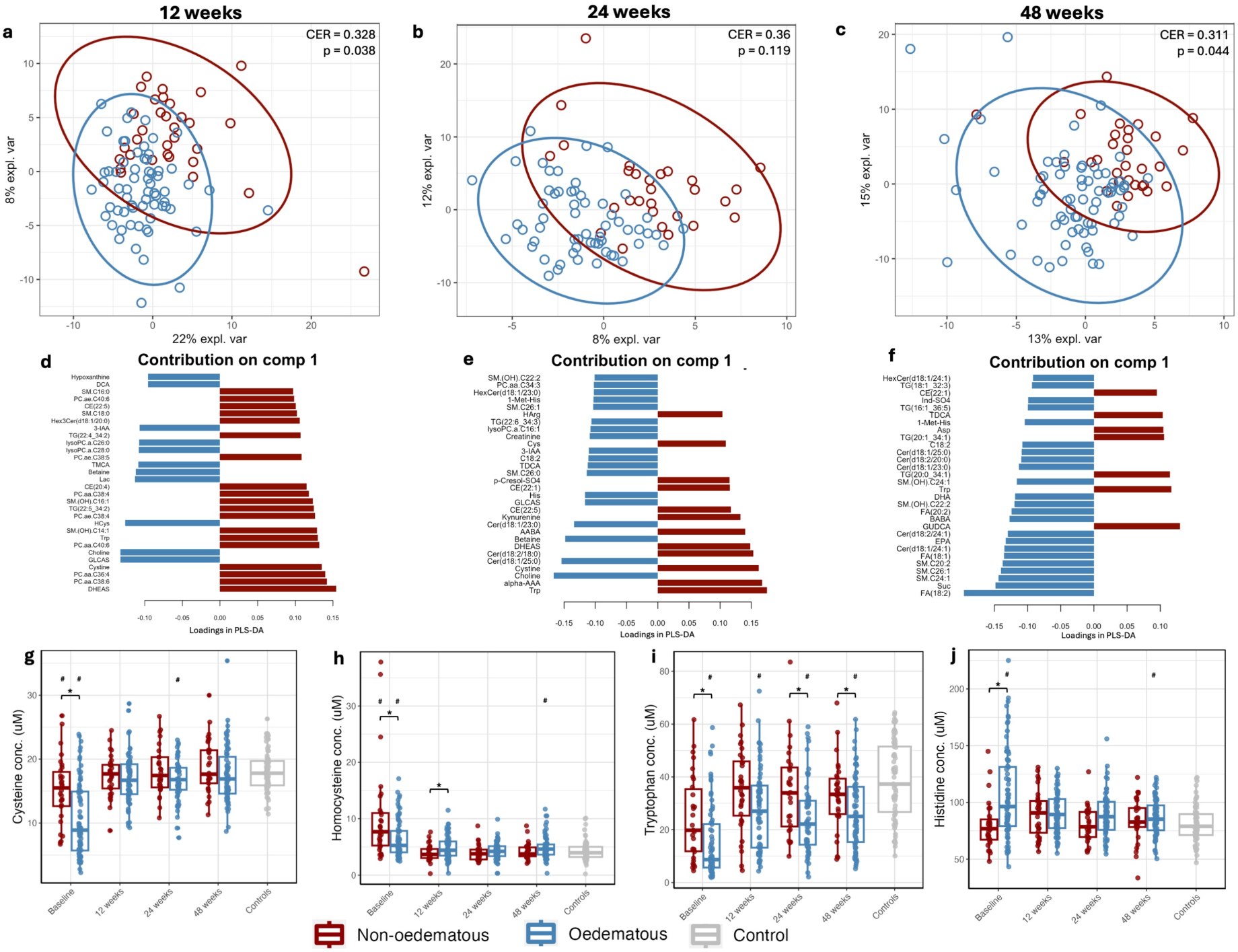
Plasma metabolomes remain distinct between O-SAM and NO-SAM up to 48 weeks after hospital discharge. (a-c) PLS-DA models of O-SAM versus NO-SAM at follow-up visits including model results from permutation test (p value and classification error rate (CER)). (d-f) top metabolites contributing to model separation based on model loadings scores. (d-i) Concentrations of selected metabolites in O-SAM, NO-SAM and ANC during follow-up visits (*indicates significant model comparing O-SAM versus NO-SAM within a follow-up visit; ^#^indicates significant result for a model comparing O-SAM or NO-SAM with ANC).

## DISCUSSION

The pathophysiology of O-SAM remains elusive and no specific treatment strategies are available that differ from the management of NO-SAM despite differences in mortality and clinical outcomes for these two phenotypes of SAM. In this large, multi-site, observational study of complicated SAM in sub-Saharan Africa, we found that children with O-SAM had differences in one-carbon metabolism versus NO-SAM characterized by depleted circulating cysteine and homocysteine. Disturbance of one-carbon metabolism has been reported previously in O-SAM, but it has been unclear whether this is due to dietary deficiency of one-carbon metabolites in the form of sulfur amino acids, or other mechanisms. We found that the gut microbiome of children with O-SAM differed significantly from NO-SAM despite the controlled hospital setting and identical treatment protocols. Intriguingly, these microbial differences were characterized by enrichment of bacterial species involved in the metabolism of cysteine into H_2_S, namely *Lancefieldella parvula,* in addition to the increased abundance of metagenomic pathways and genes involved in methylation and one-carbon metabolism, amongst other pathways. These particular microbiome changes were not evident during 1 year of post-discharge follow-up; however, children with NO-SAM displayed delayed recovery of their gut microbiomes relative to O-SAM and ANC throughout this period. Although differences in cysteine and homocysteine abundances were attenuated between SAM groups throughout follow-up, both SAM groups displayed distinct metabolic phenotypes relative to each other and ANC for up to 48 weeks post-discharge. Collectively, these data extend previous results by other groups showing differences in one-carbon metabolism between O-SAM and NO-SAM that persist for many months despite nutritional rehabilitation, and we show for the first time that these differences may be related to distinct signatures in the gut microbiome.

In the HOPE-SAM cohort, children with O-SAM had two-fold higher mortality than those with NO-SAM during hospitalisation. However, mortality and hospital readmission were higher and nutritional recovery was lower in NO-SAM throughout 48 weeks of follow-up. The pathophysiological mechanisms contributing to these differing clinical outcomes are uncertain. The dogma that O-SAM is caused by protein insufficiency is challenged by data showing that overall protein intake does not differ between children with O-SAM and NO-SAM (26, 27) and that identical twins from the same home consuming the same diet can become discordant for O-SAM (20). Furthermore, O-SAM can be resolved even whilst consuming a protein-deficient diet (28, 29). However, more recent evidence suggests that intake of sulfur amino acids may be lower in regions with high prevalence of O-SAM versus regions with low prevalence (14). Low sulfur amino acid intake therefore, may be a necessary, but not sufficient, factor contributing to risk of O-SAM (12). Metabolic flux studies show that children with O-SAM versus NO-SAM display more efficient use and less oxidation of dietary cysteine and slower endogenous production of methionine, suggesting a higher dietary requirement for both amino acids in O-SAM (30, 31); however, the reasons for this difference in amino acid requirements are unknown. Both historic and recent metabolic phenotyping studies consistently show lower amounts of cysteine, homocysteine and methionine in the plasma and urine of children with O-SAM, whilst other metabolites involved in one-carbon metabolism are elevated (11, 12). Cysteine-rich proteins are likely lost at a higher rate due to greater atrophy of the intestinal mucus layer and damage to skin and hair in O-SAM versus NO-SAM. Indeed, some evidence suggests that children with O-SAM display greater gut mucosal atrophy (32, 33), which is a site of mucosal proteins rich in cysteine. In the gut, this intestinal dysfunction, partly driven by HSPG and sulphated GAG depletion (13), leads to protein leakage and hypoalbuminemia. The distinct intestinal changes that are observed in O-SAM suggest that the intestinal mucosal barrier may act as a potential target for intervention to reverse the metabolic disturbances related to sulfur amino acid and one-carbon metabolism and improve O-SAM clinical outcomes.

The gut is a major site of sulfur metabolism and it is estimated that 25% of whole body transsulfuration occurs in the gut where 20-25% of sulfur compounds, including methionine and cysteine, are metabolized to H_2_S (34). Preclinical evidence supports the importance of the gut microbiome for sulfur amino acid metabolism and antioxidant capacity in the host, whereby antibiotic-induced ablation of the gut microbiota reduces antioxidant reactive sulfur species in the circulation (35). Absence of a microbiome, such as in germ-free mice and pigs, alters one-carbon metabolism; increased methionine and reduced homocysteine concentrations are observed in the large intestine and in serum (36). However, the association between gut microbial function and host one-carbon metabolism remains underexplored in human clinical studies. Importantly, cysteine is a precursor to the antioxidant glutathione, which is consistently depleted in O-SAM (37). Methionine, another sulfur amino acid, is also converted to glutathione via *S*-adenosyl-L-methionine. Cysteine supplementation improves the glutathione synthesis rate in children with O-SAM (37), but in a randomized trial of a mixture of *N*-acetylcysteine and other antioxidants, there was no effect in prevention of O-SAM (10). The reason for the negative outcome of this trial is unclear, but it suggests that either antioxidant depletion is a consequence rather than a cause of O-SAM or that higher antioxidant doses could have a preventative effect on O-SAM. O-SAM has long been considered a condition of oxidative stress, whereby depletion of antioxidants leads to accumulation of reactive oxygen species driving inflammation and fluid leakage, exacerbating SAM. However, it has to date been unclear what is driving this oxidative stress.

*L. parvula* was one of only three species present at significantly higher relative abundance in O-SAM versus both NO-SAM and ANC following multivariate regression and FDR testing. *L. parvula*, in addition to *V. parvula* which was elevated but did not meet FDR thresholds for significance, produce H_2_S from cysteine. Other microbial H_2_S producers, namely *Bilophila wadsworthia* have previously been observed to be overrepresented in O-SAM stool microbiomes transplanted into germ-free mice, when compared to transplanted microbiomes from ANC (20). H_2_S plays a regulatory role in intestinal barrier function by acting as an energy source for colonocytes (38), however excess intestinal H_2_S that exceeds capacity for oxidation by colonocytes disrupts the mucus layer. H_2_S-producing species are also overrepresented in the microbiome of children with inflammatory bowel disease (IBD), who experience intestinal inflammation and barrier dysfunction (39). *L. parvula* is central to the network of microbiome differences in children with IBD and induces colitis when administered to susceptible mice. Histological studies of the intestinal epithelium in O-SAM demonstrate mucus disruption that could plausibly be induced in part by H_2_S and associated H_2_S-producing members of the microbiome (32). Evidence from children with stunting also found *Veillonella* species to be strongly associated with the small intestinal inflammation caused by environmental enteric dysfunction (EED) and these EED phenotypes could be recapitulated in mice through activity of *V. parvula* and other species (40).

Other species and metabolites also contributed to differences in the gut microbiome and metabolic phenotypes in this cohort. Prevotella sp 885 was elevated in O-SAM versus NO-SAM during baseline and during follow-up whereas other Prevotella species, notably *P. copri* clades, showed similar patterns during follow-up. A depletion of Prevotella has been observed previously in NO-SAM (18) and is critical in mediating response to nutritional interventions (41). As a key species in non-industrialized microbiomes (42), its depletion may reflect the more wasted state of children with NO-SAM. A majority of differentially abundant metabolites were elevated in O-SAM versus NO-SAM which included several phosphatidylcholines and a ceramide. This contrasts with previous data showing global metabolite depletion in O-SAM, including depletion of phosphatidylcholines. However, our data are consistent with previous data showing depletion of one-carbon metabolites cysteine and methionine (12) in addition to tryptophan and kynurenine (11). It has previously been hypothesized that liver steatosis in O-SAM drives depleted circulating phosphatidylcholines through disrupted lipid metabolism and transport, but we did not observe this here.

During follow-up of participants, we observed continued differences in microbiome composition between O-SAM and NO-SAM following discharge from hospital. These differences reduced over time, with only one species differing significantly between groups by 48 weeks post-discharge. Children with NO-SAM exhibited delayed recovery of microbiome diversity relative to ANC, which was not observed in O-SAM. These results correlate with our observations in the larger HOPE-SAM cohort that children hospitalised with NO-SAM had higher mortality and hospital readmission and poorer nutritional recovery throughout the follow-up period. NO-SAM also has a longer onset period versus the acute onset of O-SAM, providing rationale for the longer clinical, nutritional and microbiome recovery in NO-SAM. Some of the microbiome and metabolic differences in O-SAM such as the elevated abundance of *L. parvula* and the depletion of cysteine, were only observed during hospitalisation and not during follow-up, suggesting that the acute disturbances observed in cysteine metabolism in O-SAM may be microbiome-driven pathways, which was partly validated through modest correlations between plasma cysteine and metagenomic pathways involved in sulfur amino acid metabolism. On the other hand, the depletion of tryptophan in O-SAM versus NO-SAM was sustained throughout 48 weeks of follow-up even after gut microbiome profiles had normalized, suggesting that disruption to tryptophan metabolism in O-SAM may be driven by host metabolic pathways independently of the gut microbiome.

A growing body of evidence supports the influence of the gut microbiome in recovery from SAM and the potential value of microbiome-directed complementary foods (MDCF). Extensive work recapitulating features of SAM in germ-free mice led to the production of an MDCF that enhanced growth recovery in children with moderate acute malnutrition (MAM) and SAM to a greater extent than standard RUTF (21, 22). The data reported here support the central role of the microbiome in the pathophysiology of SAM, whilst providing a number of novel insights. Firstly, in contrast to studies focused on community cases of uncomplicated SAM, the current study enrolled children with complicated SAM – a more critical condition with multiple comorbidities and high mortality, for which a novel microbiome-targeted therapy is urgently needed. Secondly, the use of shotgun metagenomic sequencing provides greater insight into the metagenomic pathways through which the gut microbiome may contribute to metabolic and clinical differences in O-SAM, extending previous work using 16S sequencing. Finally, the paired ‘omics techniques, which have previously only been used in isolation in smaller cohorts, allow for a comprehensive characterization of O-SAM and identify interactions potentially driven by the microbiome that involve sulfur-containing amino acids, one-carbon metabolism and downstream antioxidant capacity.

This study has a number of limitations. Recruitment into the HOPE-SAM study only occurred after admission to hospital and clinical stabilisation of the child, resulting in a median 3 days of hospitalisation before baseline sample collection. It is possible that the sickest children died prior to study enrolment. Furthermore, antibiotic treatment is part of the WHO treatment protocol for SAM and would be expected to have a strong influence on gut microbiome and metabolomic profiles prior to baseline stool sampling. However, children with both O-SAM and NO-SAM were treated similarly with antibiotics, thereby reducing the influence of confounding by antibiotic therapy in the comparison of the SAM groups. Secondly, stool and plasma samples were often difficult to collect due to the clinical state of participants. We did not perform any sample selection for stool microbiome analysis and instead used every available stool sample, which may have led to bias. Due to biosafety protocols for HIV-positive plasma samples, shipping restrictions and insufficient funding, analysis of plasma metabolic phenotypes was limited to samples from HIV-negative children from the two Zimbabwe study sites which also may have biased our results. However, we compared baseline characteristics for those enrolled in the enteropathy sub-study who were included in our analysis versus those not included and found overall comparable characteristics apart from significantly larger MUAC and lower mortality in included children, suggesting we did not capture some of the more severe clinical cases. For dataset integration analyses, only 86 participants had paired stool microbiome and plasma metabolome data available at baseline, which limited our statistical power to detect small effects.

## Conclusions

Deep multi-omic analysis identified strong associations between gut microbiome profiles, metabolic phenotypes and oedema status in children hospitalised with SAM that suggest a potential influence of the gut microbiome on disturbed one-carbon metabolism in O-SAM. Impairment of anti-oxidant capabilities, driven by the microbiome, is likely to dampen the resilience of children with O-SAM in the face of infections or nutritional deficiencies and contribute to their poorer clinical outcomes. Metabolic and microbiome derangement persists in both SAM phenotypes during convalescence which may contribute to poor long-term growth and clinical outcomes. These data suggest that adjunctive treatments that correct gut microbiome disturbances may help to correct metabolic imbalances for O-SAM and the associated poor clinical outcomes.

## METHODS

### Study participants

This was a sub-study within the Health Outcomes, Pathogenesis and Epidemiology of Severe Acute Malnutrition (HOPE-SAM) study, which has been described in detail elsewhere (25). The HOPE-SAM protocol, standard operating procedures, and case report forms are available at https://osf.io/29uaw/. Briefly, HOPE-SAM was a longitudinal observational cohort study of children hospitalised for SAM at three tertiary referral hospitals in Zambia and Zimbabwe between August 2016 and March 2018. Eligible children were younger than 60 months of age and admitted to a medical ward with complicated SAM, defined as WHZ < -3 or mid-upper-arm circumference (MUAC) <115mm and/or the presence of bilateral pitting oedema (in children above six months of age), according to WHO criteria. Children with malignancy, who died before study enrolment, or whose caregiver was not willing to learn the child’s HIV status were not included in the study. An enteropathy sub-study enrolled children aged 6-59 months, stratified by HIV status, to more intensive longitudinal specimen collection during follow-up; because the goal was to evaluate biomarkers of enteropathy, children with chronic gastrointestinal diseases were excluded.

Caregivers were sensitised to the study upon admission of their child to hospital for in-patient treatment. Following initial stabilization of children, potential participants were screened and enrolled (baseline visit occurred a median 3 days after hospital admission). Due to this gap in time between admission and recruitment, in some cases, oedema had resolved between admission and baseline sample collection. Oedema status upon admission was used to classify O-SAM and NO-SAM groups. Upon enrolment, baseline demographic and clinical information was collected on case report forms. Baseline samples were collected following recruitment to the study. In-patient care was provided using WHO country-adapted guidelines for management of complicated SAM. Children underwent a clinical examination and anthropometric measurement, including weight, height, MUAC, and head circumference, and history of illness and hospital readmission were obtained. Stool and plasma samples were collected from sub-study children at baseline, hospital discharge, 12, 24, and 48-week post-discharge visits.

To provide normative biomarker data for the local population, single samples of blood and stool were collected from adequately-nourished control (ANC) children receiving in-patient or outpatient care at each site. Controls were aged 6-59 months, with WHZ>-1, no acute illness or current infections, and known HIV status. Controls were matched within age bands (6-11 months, 12-23 months, 24-59 months) and by HIV status to enrolled sub-study children with SAM.

### Ethics

Research ethics approval for the study was obtained from the University of Zambia Biomedical Research Ethics Committee (010-02-16), and the Medical Research Council of Zimbabwe (MRCZ/A/2044). The ethics committee of Queen Mary University of London provided an advisory review. Written informed consent from caregivers was obtained in local languages.

### Sample collection and storage

Blood samples were collected by research nurses in endotoxin-free EDTA tubes during hospitalisation and at follow-up visits at the hospital sites. Blood samples were transported in cool boxes to the research laboratories where plasma was isolated and stored at -80°C within the same day. Stool samples were collected from participants in sterile containers without preservation buffer and kept cool until transfer to the laboratory for aliquoting and freezing at -80°C within the same day. Children with nasogastric tubes during hospitalisation had 3-5ml gastric fluid collected in sterile containers without preservation buffer using a sterile syringe; samples were kept cool until transfer to the laboratory for aliquoting and freezing at -80°C within the same day. Stool and gastric fluid aliquots for microbiome sequencing were shipped on dry ice to the British Columbia Centre for Disease Control, Vancouver, Canada for processing.

### Sample selection

All available stool samples collected in the study were included for microbiome analysis. Due to constraints with biosafety protocols, shipping restrictions and funding, we selected plasma samples for metabolomics that were (i) HIV negative; (ii) from the two Zimbabwean sites only; and (iii) had a paired stool and/or urine sample (urine analysis not reported here). All remaining children <18 months old at baseline who met the first two inclusion criteria but did not have a paired urine/stool sample were also included in the metabolomics analysis.

### Whole metagenome shotgun sequencing

DNA was extracted from 100–200 mg of stool samples using “Protocol Q” from the International Human Microbiome Standards (http://www.microbiome-standards.org/) as reported previously (43). DNA quantity was assessed by fluorometry (Qubit) and quality confirmed by spectrophotometry (SimpliNano). 1 µg DNA was subsequently used as input for metagenomic sequencing library preparation using the Illumina TruSeq PCR-free library preparation protocol, using custom end-repair, adenylation and ligation enzyme premixes (New England Biolabs). The concentration and size of constructed libraries were assessed by qPCR and by TapeStation (Agilent). DNA-free negative controls and positive controls (ZymoBIOMICS D6300) were included in all DNA extraction and library preparation steps. Libraries were pooled in random batches of 48 samples including one negative control. Whole metagenome sequencing was performed with 125-nucleotide paired-end reads using the Illumina HiSeqX platform at Canada’s Michael Smith Genome Sciences Centre, Vancouver, Canada.

### Bioinformatics

Quality control and bioinformatic processing of raw sequencing data was conducted using the publicly available KneadData (https://huttenhower.sph.harvard.edu/kneaddata/). Sequenced reads were trimmed of adaptors and filtered to remove low-quality, short (<70% raw read length), and duplicate reads, as well as those of human, other animal or plant origin, with default settings. Species composition was determined by identifying clade-specific markers from reads using MetaPhlAn4 with default settings. Relative abundance estimates were obtained from known assigned reads. We applied a minimum threshold of >0.1% relative abundance and ≥5% prevalence for all detected species. Metabolic pathway composition was determined using HUMAnN3 with default settings against the UniRef90 database. Enzyme commission (EC) and pathway abundance estimates were normalized using reads per kilobase per million mapped reads (RPKM) and then re-normalized to relative abundance. We applied a minimum relative abundance threshold of 3 × 10^−7^% and ≥5% prevalence for all metagenomic pathways.

### Targeted metabolic phenotyping

Plasma metabolic profiles were measured on a Waters Acquity Premier ultra-performance liquid chromatography system coupled to a Xevo TQ-XS mass spectrometer using the Biocrates MxP Quant 500 kit (Biocrates Life Sciences, Innsbruck, Austria). This kit incorporates internal standards that are used for metabolite identification and quality control samples to assess reproducibility. This data was acquired following the manufacturer’s instructions. Quantification was achieved using a seven-point standard curve of internal standards. Briefly, 10 μl of thawed sample was added to a 96-well plate containing inserts of internal standards supplied by Biocrates and dried under nitrogen. Samples were incubated with 5 % phenyl isothiocyanate (PITC), dried, shaken followed by the addition of 5 mM ammonium acetate in methanol. Samples were transferred to a separate 96-well plate and diluted with either pure water or FIA solvent. Metabolites were measured in positive and negative ionization mode. The LC-MS/MS method involved the injection of 5 µl of extract into the Biocrates MxP Quant 500 kit UHPLC column at 50 °C using a 5.8 min solvent gradient with 0.2 % formic acid in water and 0.2 % formic acid in acetonitrile. The FIA-MS/MS method involved the injection of 20 µl of extracts. The raw data from the UPLC-MS and FIA analyses were processed using the Biocrates MetIDQ software (version Oxygen-DB110-302305) in accordance with the manufacturer’s protocol. Batch correction was performed to minimize technical variation across the analytical run using standard quality control samples dispersed throughout each plate.

### Statistical analysis

All data were analysed using R (v.4.3.2). Microbiome data were handled using the phyloseq package (v1.46). Alpha diversity metrics were calculated using the vegan package (v2.6.8). Differences in alpha diversity were assessed by univariate and multivariate linear regression in cross-sectional analysis at each time point. Beta-diversity was estimated using the Bray-Curtis dissimilarity index and analysed by permutation testing in the EnvFit package at baseline. For comparison between O-SAM, NO-SAM and ANC at follow-up visits, permutational analysis of variance (PERMANOVA) with Bray-Curtis distances was applied. Differential abundance analysis of species or functional pathways by oedema status was assessed using the MaAsLin2 (v1.16) package for multiple regression (68) where default arguments were supplied and applying significance thresholds of p<0.05, q<0.3. Three covariates were chosen for adjustment in diversity analyses and MaAsLin2 regression models; these included age, current breastfeeding status (binary variable based on the question: “Is this child currently being breastfed?”), and HIV status. These covariates were chosen following construction of a directed acyclic graph (DAG; Fig. S8). Adjustment for multiple comparisons was performed using the Benjamini-Hochberg false discovery rate (FDR).

To test for differences in plasma metabolites, univariate and multivariate linear regression analyses were performed, adjusting for age, sex and current breastfeeding status. Adjustment for multiple comparisons was performed using the Benjamini-Hochberg false discovery rate (FDR). PLS-DA and sPLS-DA analysis was conducted using the mixOmics package. For data integration, we used Data Integration Analysis for Biomarker discovery using Latent cOmponents (DIABLO), a multi-omic data integration method that uses sparse generalized canonical correlation analysis (sGCCA) to simultaneously identify key omics variables during the integration process and discriminate phenotypic groups. The DIABLO model included three datasets: microbiome species, microbiome pathways and plasma metabolites.

## Data Availability

The raw metagenome sequencing data generated in this study have been deposited in the European Bioinformatics Database

## ACKNOWLEDGEMENTS

We thank the children and caregivers who participated in the HOPE-SAM study and the staff at Harare Central Hospital, Parirenyatwa Hospital and the University Teaching Hospital for hosting and supporting the study team. We acknowledge the wider HOPE-SAM study team (members listed here: https://bmjopen.bmj.com/content/9/1/e023077) and Stephen Moyo (logistics), Philippa Rambanepasi (finance), Virginia Sauramba and Miyoba Chipunza (compliance), and Agatha Muyenga (administration), in particular, for their additional support for this study at Zvitambo and TROPGAN.

## FUNDING

HOPE-SAM was funded by the Medical Research Council (MR/K012711/1), Wellcome (107634/Z/15/Z to MB, 206455/Z/17/Z to RCR, and 093768/Z/10/Z and 108065/Z/15/Z to AJP), a joint award from Wellcome and the Royal Society (206225/Z/17/Z to CDB), and UNICEF Zimbabwe (ZIM/PCA201721/PD2019158). JRS is supported by *NIHR Southampton Biomedical Research Centre (NIHR203319)* and BBSRC (UKRI775; BB/W00139X/1). Funding bodies had no role in the study design, implementation, analysis and interpretation of the data.

## AUTHOR CONTRIBUTIONS

Conceptualization: RCR, AJP, ARM, BA, KN, PK, MBD

Data and specimen collection and processing: RCR, TJE, IB, NV, CD, MP, KM, SM, FF, EB, TR, DN, FDM, KZ, JRS, CDB, SR, MG,

Data management: CD, RN, BC

Data analysis and interpretation: RCR, TJE, JRS, PK, ARM, AJP

Funding acquisition: RCR, AJP, PK, MDB, BA, KJN

Project administration: AJP, RCR, PK, ARM, MBD

Supervision: AJP, ARM, PK, JRS

Writing – original draft: RCR

Writing – review & editing: All authors

## COMPETING INTERESTS

RCR is on the scientific advisory board of Biostime Institute Nutrition and Care and declares remittance from Abbott Nutrition Health Institute, Nutricia and Nestle for public conference talks outside the submitted work. All other authors declare that they have no competing interests.

## DATA AND MATERIALS AVAILABILITY

The raw metagenome sequencing data generated in this study have been deposited in the European Bioinformatics Database under accession code (*data to be uploaded after review*). Epidemiologic data files and metabolomics data are available at (*link to be provided after review*).

## SUPPLEMENTARY MATERIALS

- **Fig. S1 to S8**
- **Table. S1 to S16**

## Notes

### Clinical Protocols

https://osf.io/29uaw/

### Author Declarations

University of Zambia Biomedical Research Ethics Committee (010-02-16) and the Medical Research Council of Zimbabwe (MRCZ/A/2044) gave ethical approval for this work

