## Supplementary figures (Fig. S1 to S8) for "Gut microbiome and plasma metabolic signatures of one-carbon metabolism differentiate oedematous and non-oedematous severe acute malnutrition"

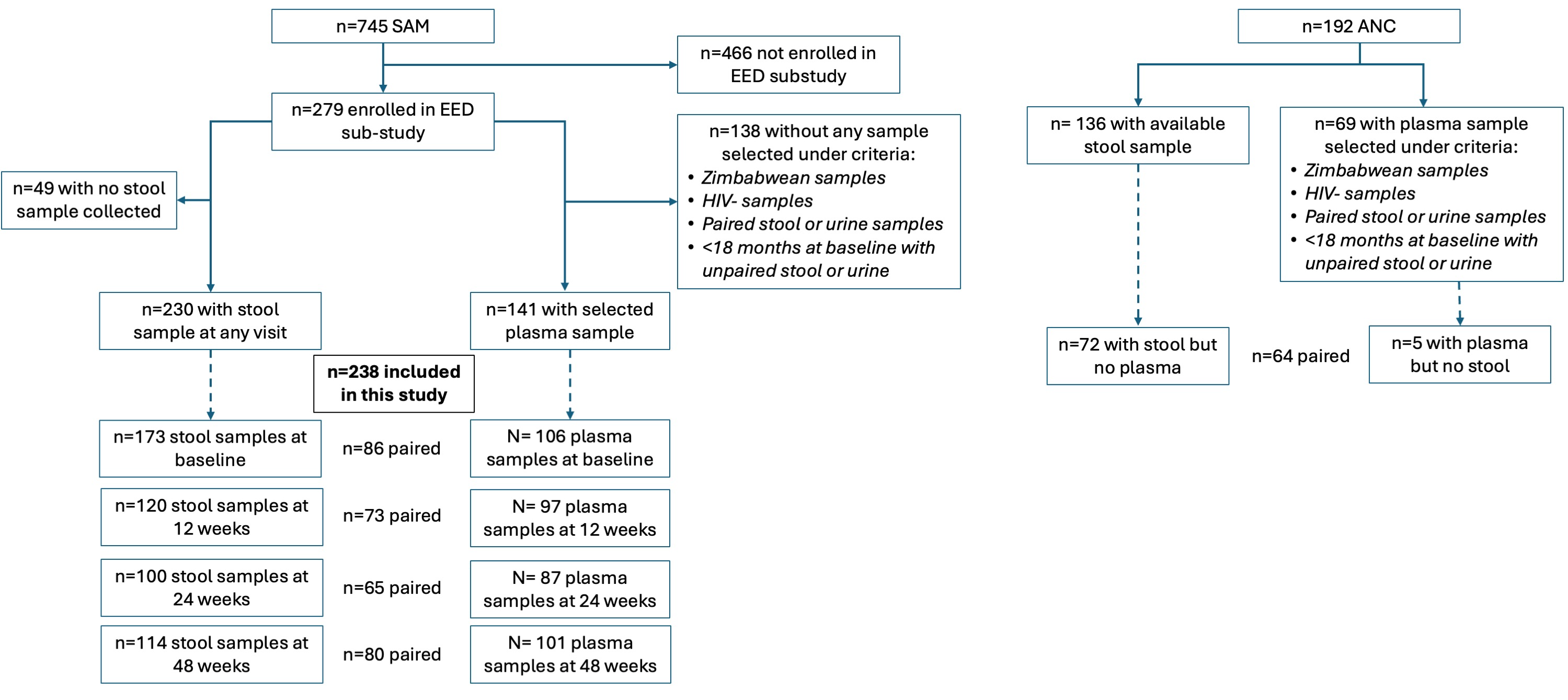

**Figure S1. Study CONSORT diagram.** CONSORT diagram of participants included and excluded from the analysis including breakdown of stool and plasma samples at each visit

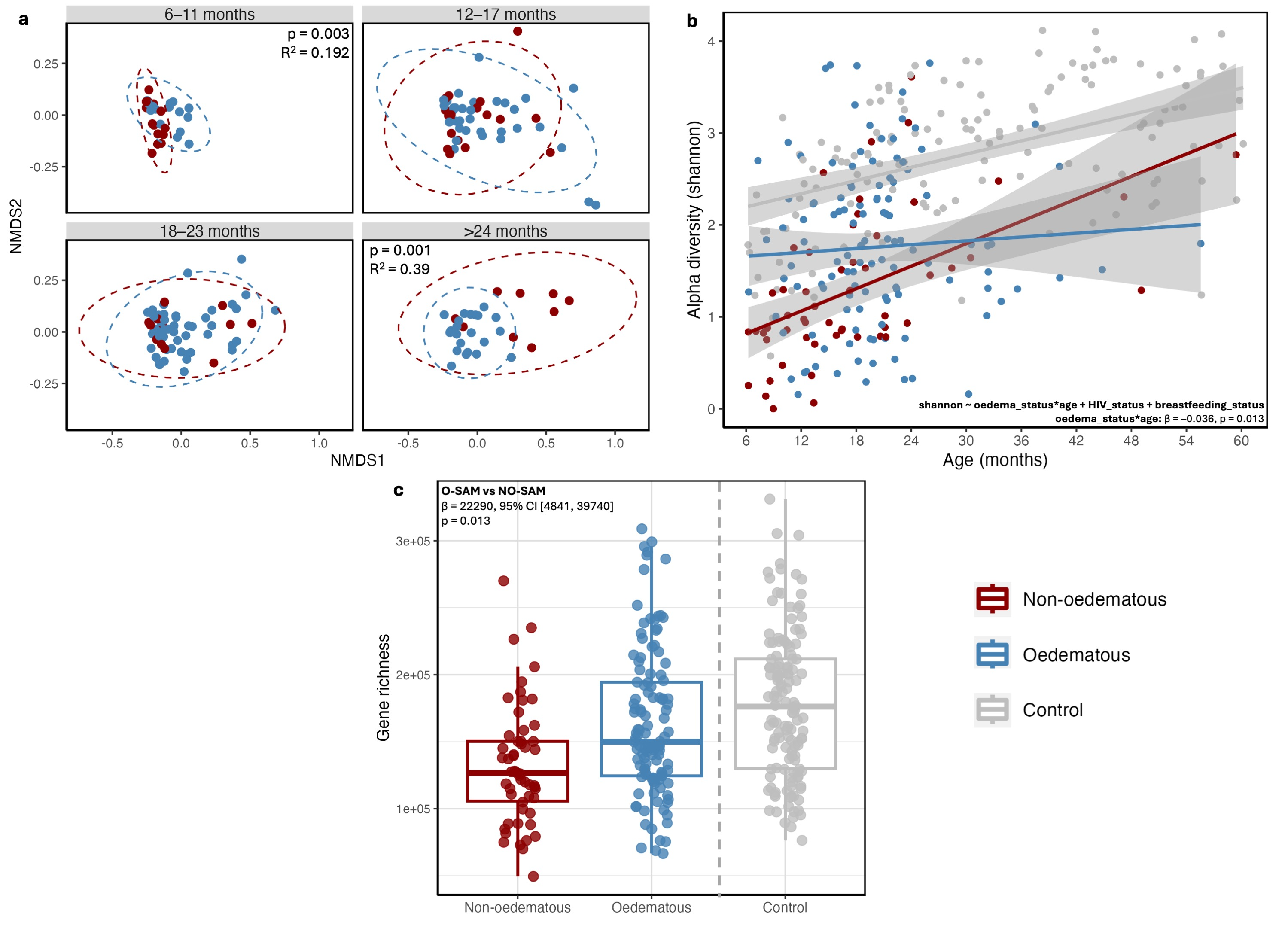

**Figure S2. Baseline microbiome taxonomic and functional diversity.** (a) NMDS ordination plots coloured by oedema status with P values and R^2^ values from EnvFit models separated by age brackets of metagenomic gene pathways. (b) Shannon diversity as a model of age and oedema status. Model results are included comparing O-SAM with NO-SAM and the interaction with age. ANC results are plotted but not included in the statistical model. (c) Alpha diversity of microbiome genes (gene richness) with results of multivariate regression model comparing O-SAM versus NO-SAM. ANC included as reference but not included in statistical model.

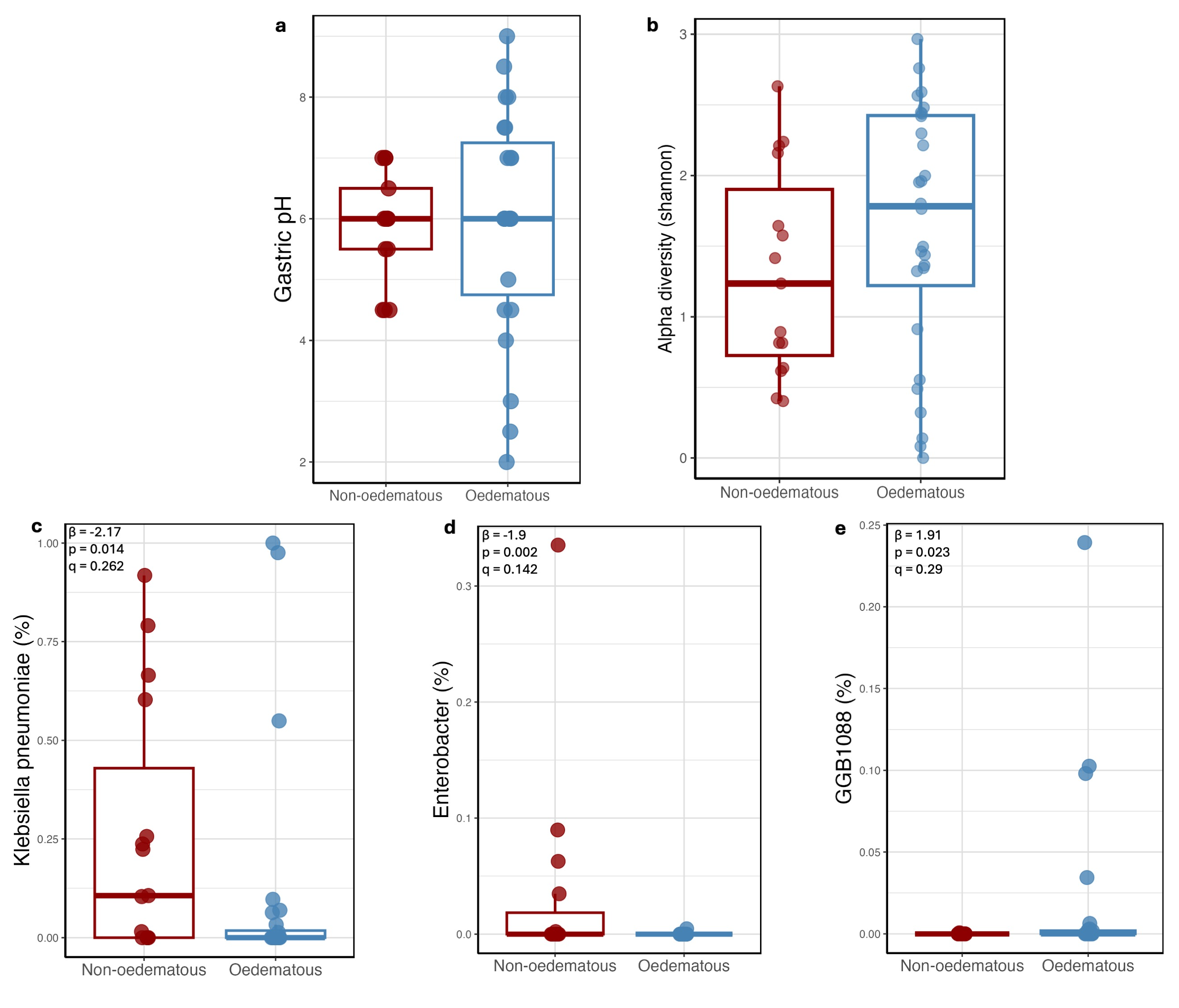

**Figure S3. Gastric fluid microbiomes during hospitalization.** (a) Gastric pH results between O-SAM and NO-SAM. (b) Alpha diversity (Shannon index) of gastric fluid microbiomes between O-SAM and NO-SAM. (c) Relative abundances and results from MaasLin2 regression for species significantly differentially abundant in the gastric fluid microbiome between O-SAM and NO-SAM.

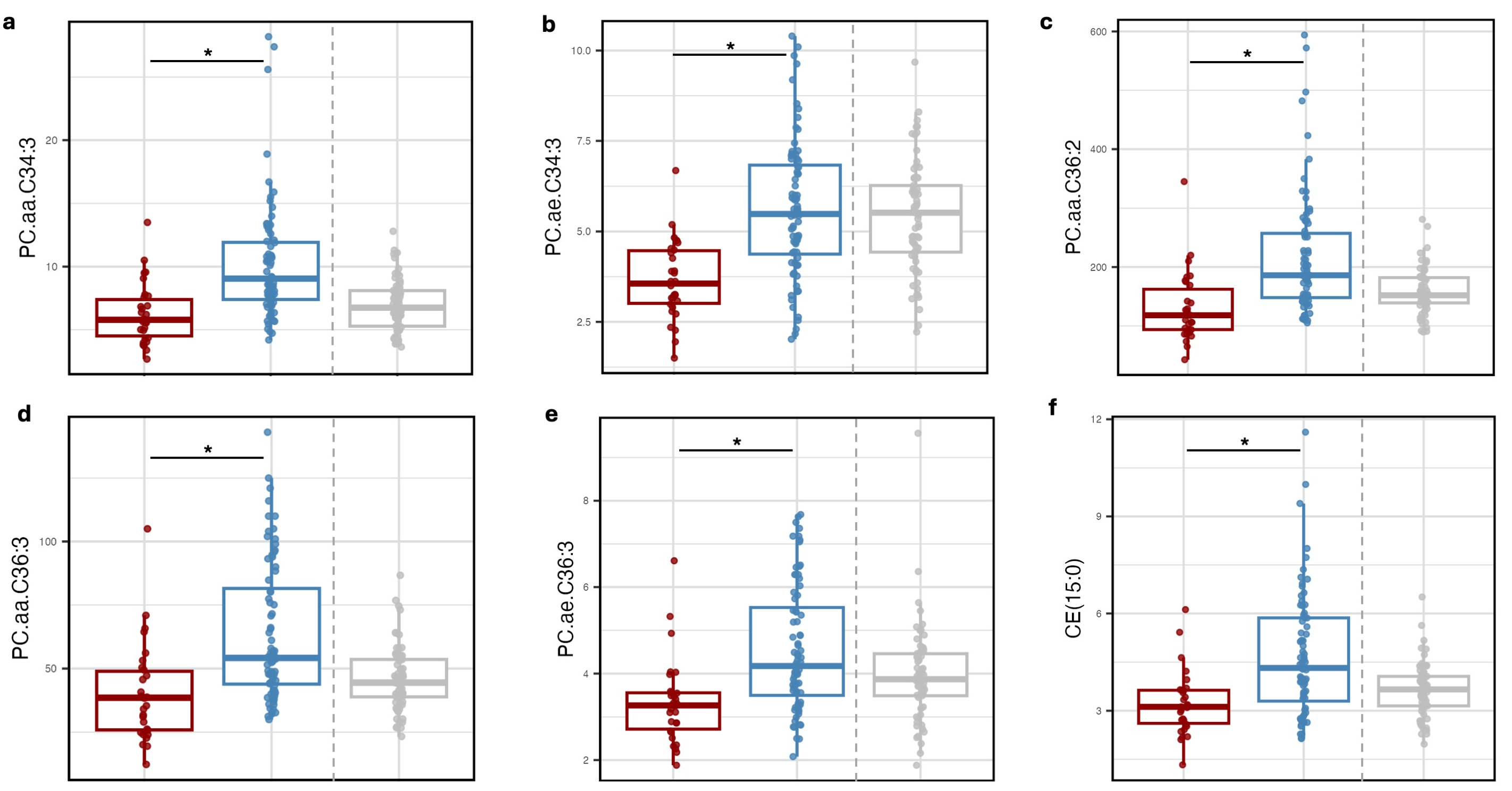

**Figure S4. Phosphatidylcholine metabolites during hospitalization.** Concentrations of the top 5 metabolites (5 phosphatidylcholines and 1 ceramide) contributing to separation in PLS-DA models of O-SAM versus NO-SAM plasma metabolomes during hospitalization (Fig. 2a). Concentrations of metabolites from ANC are plotted for reference but were not included in PLS-DA models.

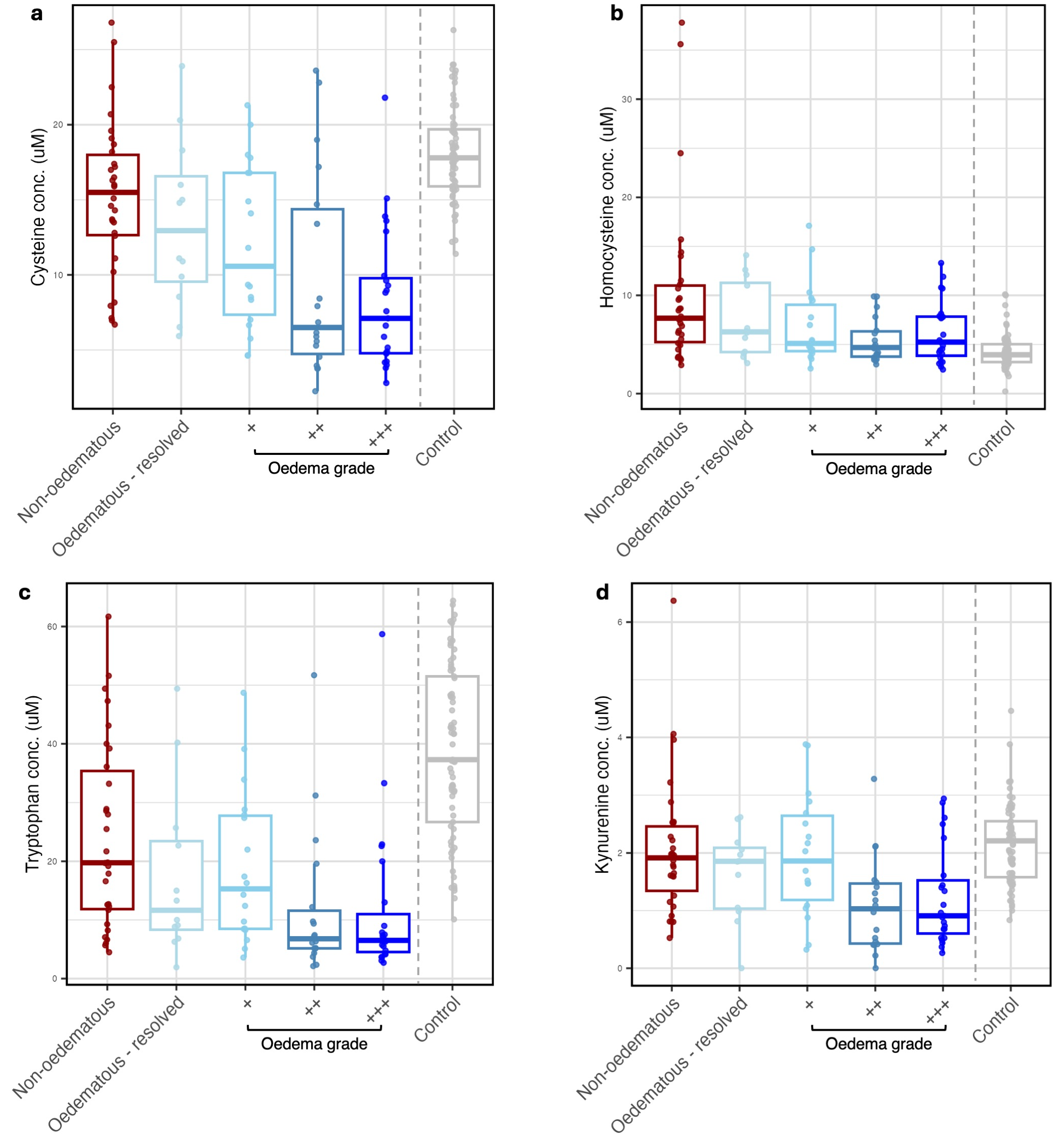

**Figure S5. Plasma metabolites depleted in O-SAM across grades of oedema severity.** (a) Cysteine, (b) Homocysteine, (c) Tryptophan and (d) Kynurenine concentrations during hospitalization across grades of oedema severity. “Oedematous resolved” refers to children admitted to hospital with O-SAM but whose oedema had resolved prior to the baseline study visit which occurred a median of 3 days after hospital admission. Oedema grades were defined as: (+) mild bilateral oedema of the feet; (++) moderate oedema of feet and lower legs or hands; (+++) generalised oedema affecting feet, legs, arms, hands and face.

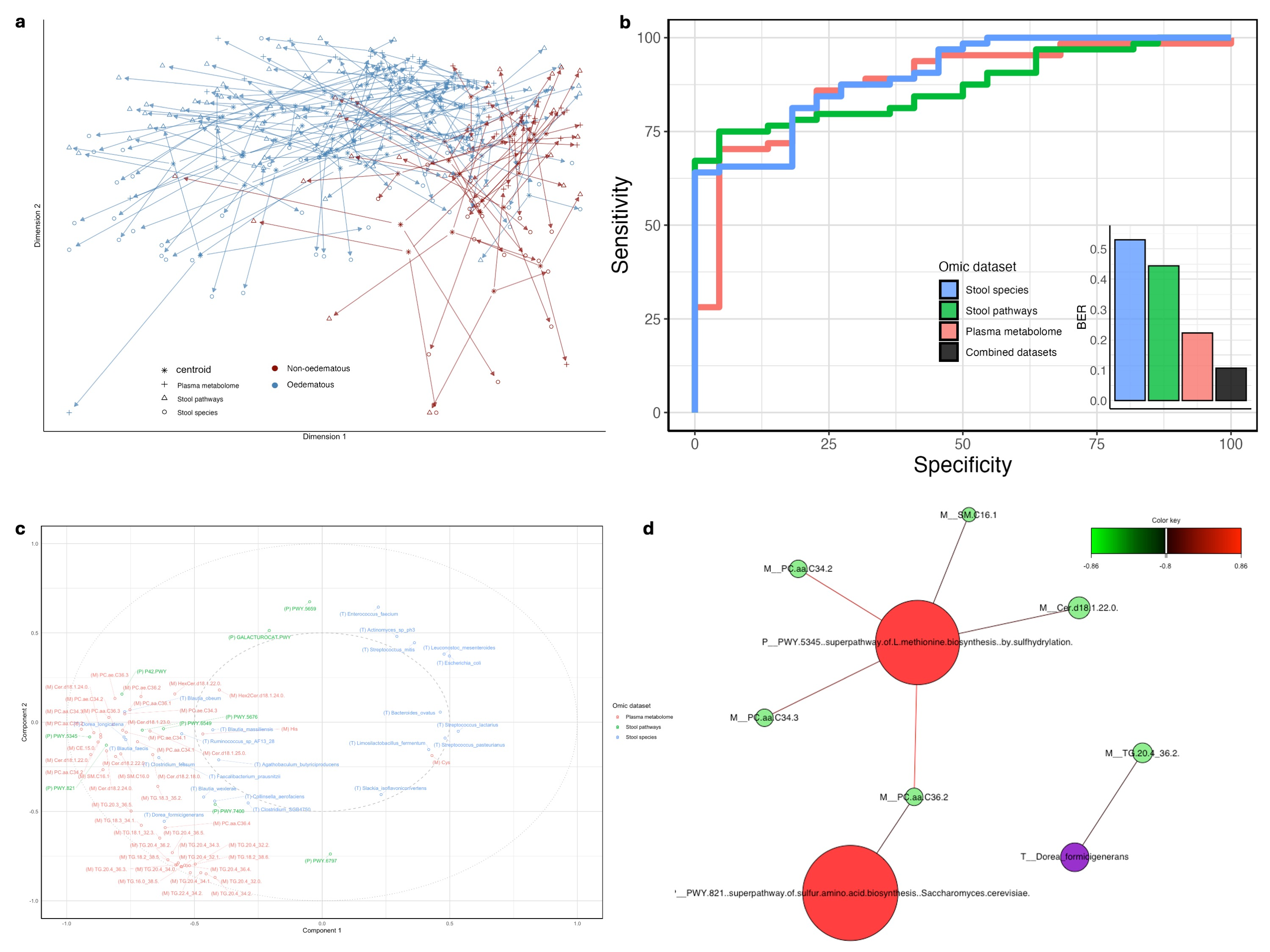

**Figure. S6. Microbiome and metabolome data integration using Data Integration Analysis for Biomarker discovery using Latent cOmponents (DIABLO).** (a) Multi-block sPLS-DA of 3 integrated datasets (microbiome species, microbiome gene pathways, plasma metabolites) shows proximity of different datasets within and between individuals and separation of O-SAM and NO-SAM groups. (b) Area under the curve (AUC) for each dataset in multiblock sPLS-DA models in DIABLO in addition to the balanced error rate (BER) for classification of O-SAM versus NO-SAM, demonstrating that the combined datasets generate the lowest error. (c) Correlation circle plots to visualise how variables within the DIABLO model relate to each other and to selected latent components (variables with correlations >0.4). (d) Network plot of interactions from DIABLO model with correlation coefficient >0.8. Node size is proportional to the number of connections (degree), and color indicates the omic data source. Edges represent pairwise correlations between variables component loadings across omics datasets.

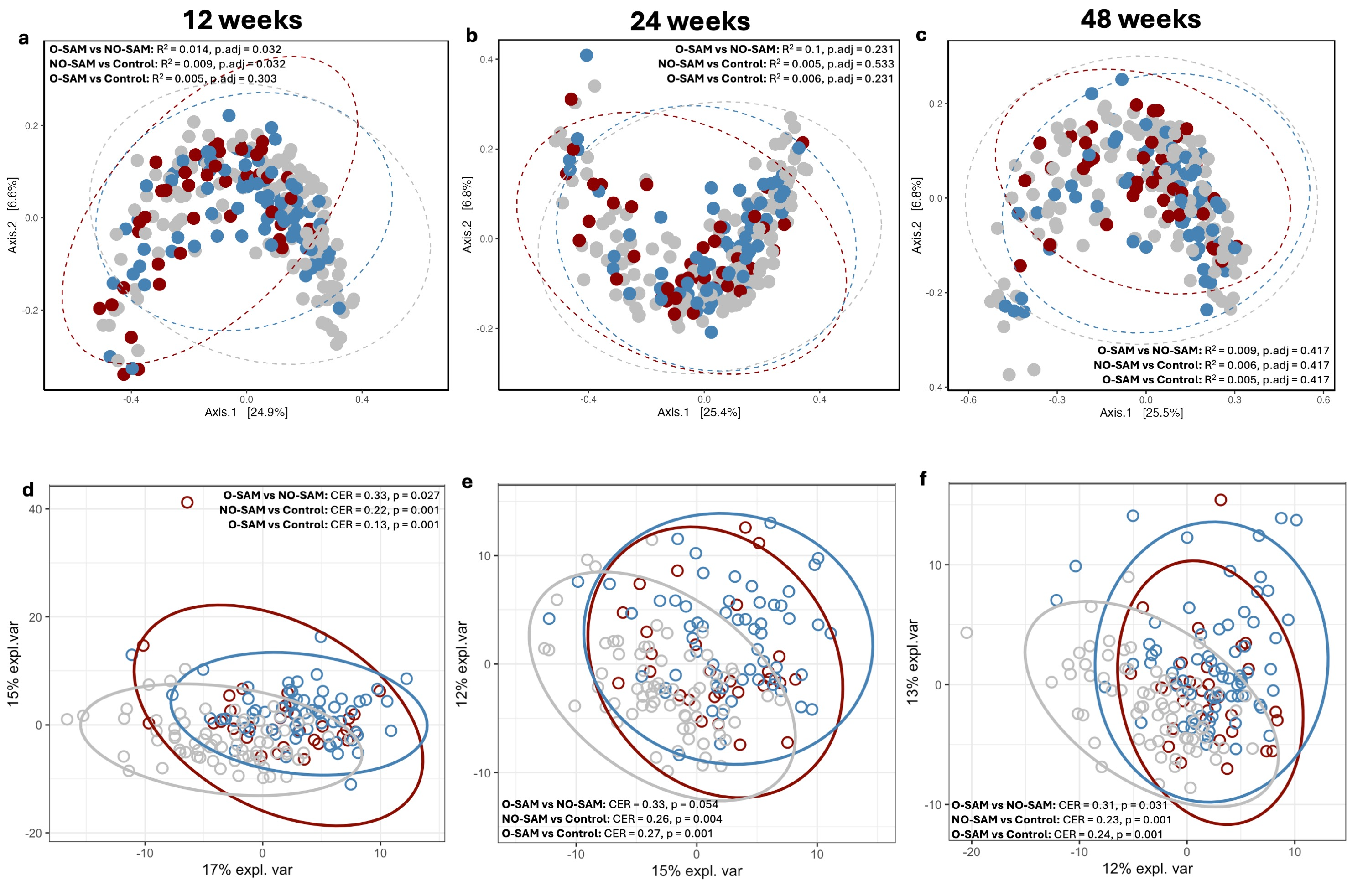

**Figure S7. Microbiome and metabolome recovery during follow-up in O-SAM and NO-SAM versus ANC.** (a-c) NMDS ordination plots coloured by oedema status with P values and R^2^ values from permutational analysis of variation (PERMANOVA) comparing NO-SAM, O-SAM and ANC. (d-f) PLS-DA models of O-SAM versus NO-SAM versus ANC at follow-up visits including model results from pairwise results of permutation tests (p value and classification error rate (CER)).

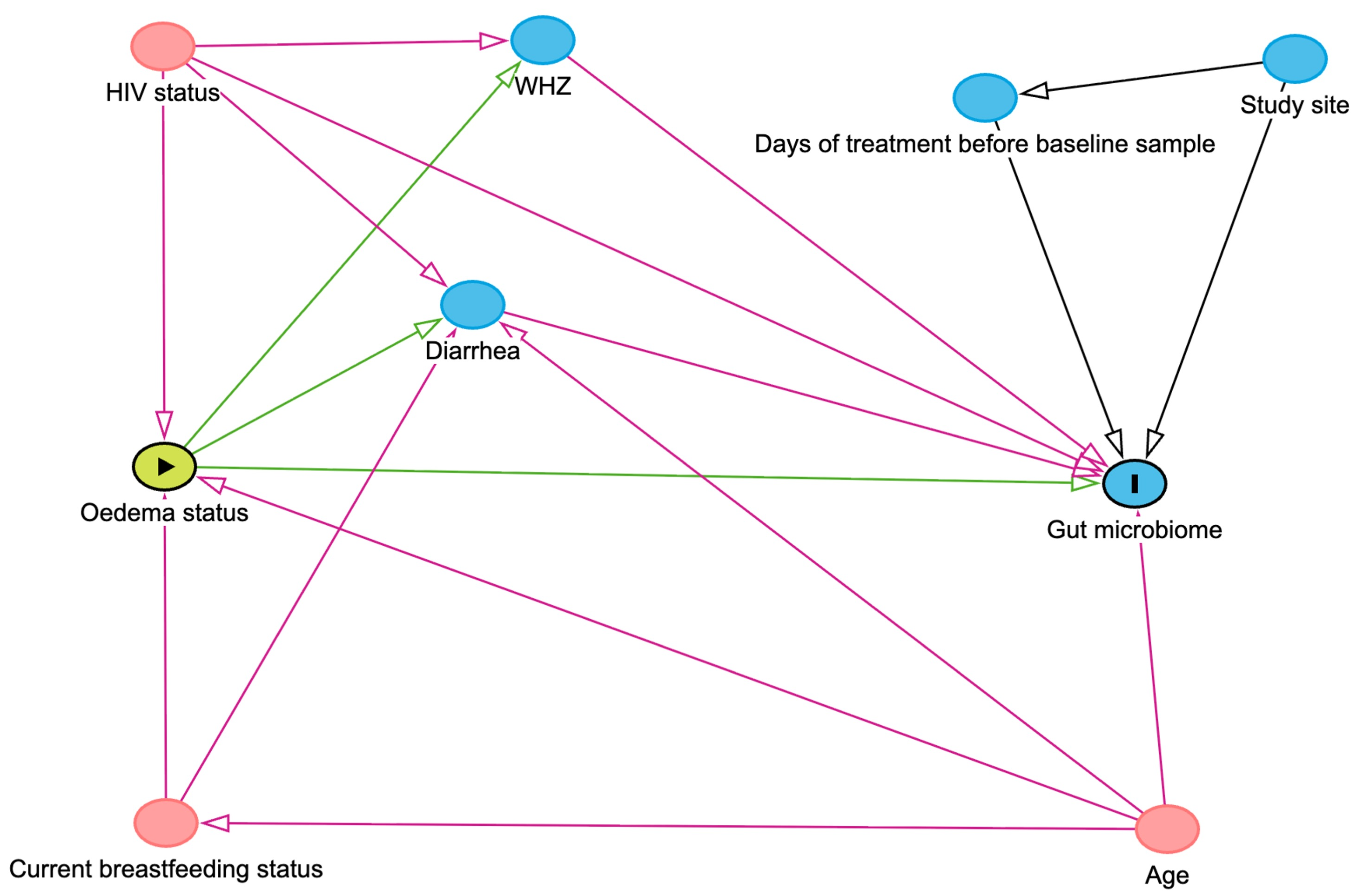

**Figure S8. Directed acyclic graph (DAG).** A DAG to assess the association between oedema status and gut microbiome identified 3 covariates (HIV status, current breastfeeding status and age) for minimally sufficient adjustment in multivariate models.
